# Ruling out SARS-CoV-2 infection using exhaled breath analysis by electronic nose in a public health setting

**DOI:** 10.1101/2021.02.14.21251712

**Authors:** Rianne de Vries, René M. Vigeveno, Simone Mulder, Niloufar Farzan, Demi R. Vintges, Jelle J. Goeman, Sylvia Bruisten, Bianca van den Corput, J.J. Miranda Geelhoed, Leo G. Visser, Mariken van der Lubben, Peter J. Sterk, Johannes C.C.M. in ’t Veen, Geert H. Groeneveld

## Abstract

**Background:** Rapid and accurate detection of SARS-CoV-2 infected individuals is crucial for taking timely measures and minimizing the risk of further SARS-CoV-2 spread. We aimed to assess the accuracy of exhaled breath analysis by electronic nose (eNose) for the discrimination between individuals with and without a SARS-CoV-2 infection.

**Methods:** This was a prospective real-world study of individuals presenting to public test facility for SARS-CoV-2 detection by molecular amplification tests (TMA or RT-PCR). After sampling of a combined throat/nasopharyngeal swab, breath profiles were obtained using a cloud-connected eNose. Data-analysis involved advanced signal processing and statistics based on independent t-tests followed by linear discriminant and ROC analysis. Data from the *training* set were tested in a *validation*, a *replication* and an *asymptomatic set*.

**Findings:** For the analysis 4510 individuals were available. In the *training* set (35 individuals with; 869 without SARS-CoV-2), the eNose sensors were combined into a composite biomarker with a ROC-AUC of 0.947 (CI:0.928-0.967). These results were confirmed in the *validation* set (0.957; CI:0.942-0.971, n=904) and externally validated in the *replication* set (0.937; CI:0.926-0.947, n=1948) and the *asymptomatic* set (0.909; CI:0.879-0.938, n=754). Selecting a cut-off value of 0.30 in the *training* set resulted in a sensitivity/specificity of 100/78, >99/84, 98/82% in the *validation, replication* and *asymptomatic set*, respectively.

**Interpretation:** eNose represents a quick and non-invasive method to reliably rule out SARS-CoV-2 infection in public health test facilities and can be used as a screening test to define who needs an additional confirmation test.

**Funding:** Ministry of Health, Welfare and Sport

**Research in context:** *Evidence before this study:* Electronic nose technology is an emerging diagnostic tool for diagnosis and phenotyping of a wide variety of diseases, including inflammatory respiratory diseases, lung cancer, and infections. As of Feb 13, 2021, our search of PubMed using keywords “COVID-19” OR “SARS-CoV-2” AND “eNose” OR “electronic nose” OR “exhaled breath analysis” yielded 4 articles (1-4) that have assessed test characteristics of electronic nose to diagnose COVID-19. In these small studies the obtained signals using sensor-based technologies, two-dimensional gas chromatography and time-of-flight mass spectrometry, or proton transfer reaction time-of-flight mass spectrometry, provided adequate discrimination between patients with and without COVID-19.

*Added value of this study:* We prospectively studied the accuracy of exhaled breath analysis by electronic nose (eNose) to diagnose or rule out a SARS-CoV-2 infection in individuals with and without symptoms presenting to a public test facility. In the *training* set with 904 individuals, the eNose sensors were combined into a composite biomarker with a ROC-AUC of 0.948. In three independent *validation* cohorts of 3606 individuals in total, eNose was able to reliably rule out SARS-CoV-2 infection in 70-75% of individuals, with a sensitivity ranging between 98-100%, and a specificity between 78-84%. No association was found between cycle thresholds values, as semi-quantitative measure of viral load, and eNose variables.

*Implications of all the available evidence:* The available findings, including those from our study, support the use of eNose technology to distinguish between individuals with and without a SARS-CoV-2 infection with high accuracy. Exhaled breath analysis by eNose represents a quick and non-invasive method to reliably rule out a SARS-CoV-2 infection in public health test facilities. The results can be made available within seconds and can therefore be used as screening instrument. The eNose can reliably rule out a SARS-CoV-2 infection, eliminating the need for additional time-consuming, stressful, and expensive diagnostic tests in the majority of individuals.

## Introduction

The ongoing spread caused by the severe acute respiratory syndrome coronavirus 2 (SARS-CoV-2) has underlined the importance of widespread, rapid and accurate testing to contain the pandemic. Despite initiation of vaccination campaigns, instant testing of all people with Coronavirus Disease 2019 (COVID-19) symptoms is recommended and propagated worldwide (5). Reverse Transcription-Polymerase Chain Reaction (RT-PCR), other molecular amplification tests such as the transcription mediated amplification (TMA), and antigen tests are currently used for detecting SARS-CoV-2 (6-8). However, performing these tests on a large-scale is time-consuming and faces other limitations, such as a shortage of personnel, equipment, reagents and other consumables (9). Delay in test results impedes timely and adequate measures and could further worsen the economic despair and the compliance to and psychological consequences of mandatory quarantine (10).

Expanding the testing capacity, analytic performance, throughput and speed of returning results using new innovative technology is urgently needed and, if successful, could contribute significantly to current efforts to curb the COVID-19 pandemic (11). Exhaled breath analysis using electronic nose (eNose) technology may be considered as a diagnostic tool during this outbreak (12, 13). Exhaled breath contains thousands of volatile organic compounds (VOCs) derived from physiological and pathophysiological processes of local and systemic origin (13, 14), which might include VOCs related to a SARS-CoV-2 infection (15). eNoses can be applied for probabilistic pattern recognition using cross-reactive sensors that capture the complete mixture of VOCs in exhaled breath (14). This emerging technology has shown potential as a point-of-care tool for diagnosis and phenotyping of a wide variety of diseases, including inflammatory respiratory diseases, lung cancer, and infections (16-18). Our hypothesis was that a SARS-CoV-2 infection is reflected by a distinct breath profile and that this profile can be identified using eNose technology. Therefore, the aim of this study was to determine the diagnostic accuracy of exhaled breath analysis by eNose for the discrimination between participants with and without a SARS-CoV-2 infection in a real-world population in the public health care domain.

## Methods

### Study design and oversight

This was a prospective, real-world study including a *training, validation, replication* and *asymptomatic set*. The study added exhaled breath analysis by eNose to the standard diagnostic procedure at the public test facility. The study protocol was approved by the Medical Ethics Committee of Leiden The Hague Delft (P20.033) and all participants provided written informed consent. The study was performed in accordance with the principles of the Declaration of Helsinki.

### Four sets of individuals

Individuals (≥18 years of age) with early symptoms suggestive of COVID-19 and/or who had been in contact with someone diagnosed with COVID-19 presented to two different Amsterdam public test facilities for a SARS-CoV-2 diagnostic test. These test centres were facilitated by the Municipal Public Health Service of the Netherlands (“GGD”). Combined throat/nasopharyngeal swabs were obtained and transported to various different molecular biological laboratories for TMA or RT-PCR. After diagnostic swab samples were obtained, individuals were asked by the study team to participate in one of four study sets between July 28 and December 27, 2020. The *training* set was specifically designed to develop the eNose diagnostic model for a SARS-CoV-2 infection, which subsequently was tested in the *validation* set. Thereafter, a *replication* set was collected in a second public test facility for external validation. Due to a change in policy in the autumn of 2020 to test more asymptomatic individuals and the lack of sufficient numbers of asymptomatics in the *training, validation* and *replication* set, a fourth independent set was collected. This *asymptomatic* set consists of individuals that visited the test facility after being in close contact with someone diagnosed with COVID-19 and did not present with new signs and symptoms of SARS-CoV-2 infection.

### Study procedures and measurements

When presenting to the test facility, all individuals underwent a combined throat/nasopharyngeal swab followed by the assessment of exhaled air using eNose technology. Demographic and clinical characteristics of the participants, e.g. age, sex, Body Mass Index (BMI), symptoms, and co-morbidities, were collected using an online clinical questionnaire. Patient-reported fever was recorded and was defined as having a body temperature >37.8 °C. Information regarding the presence and severity of cough and dyspnea was collected using Visual Analogue Scales (VAS).

In the *training* and *validation* set, the study participants were contacted by telephone by the study team after 30 days to obtain information regarding additional SARS-CoV-2 diagnostic tests and survival. If study participants were lost to follow up, information about survival was obtained from the Dutch Central Bureau for Statistics (CBS) at which mortality is registered. Due to the limited laboratory capacity, swab samples were distributed over various different laboratories. Each of these laboratories was certified for routine SARS-CoV-2 diagnostic testing using validated molecular diagnostic tests based on either TMA (only for *training* and *validation* set) or RT-PCR. This primary test result was used for the diagnostic classification. If TMA was performed initially, available stored samples from positive participants were retested by RT-PCR to obtain cycle thresholds (Ct). Ct values were used as semi-quantitative measure of viral load and were used for comparison with the eNose results.

### Diagnostic classification

In this study we used the following criteria for a definite SARS-CoV-2 infection: a positive TMA or RT-PCR at baseline and/or (only in *training* and *validation* set) a positive TMA or RT-PCR ≤ 7 days after inclusion.

### Exhaled breath analysis

Exhaled breath analysis was performed using 1) a cloud-connected eNose (SpiroNose® - Breathomix, Leiden, The Netherlands) to detect the complete mixture of VOCs in exhaled breath, 2) a gateway to securely send the eNose sensor signals to an online server and 3) an online analysis platform for real-time and automated analysis (17, 19). The SpiroNose consists of 7 different cross-reactive metal oxide semiconductor (MOS) sensors. These sensors are present in duplicate on the inside (exhaled breath measurement) and the outside (ambient air measurement) of the SpiroNose. The combination of sensor signals generates an individual breath profile, reflecting the collective VOC composition in exhaled breath. An in-depth description of the verification of the sensor stability was previously published (17, 19). Details of the complete measurement set up are provided in Figure S1 (see online supplement).

All participants were instructed to drink half a glass of water before starting the measurement. Subsequently, they performed five tidal breaths followed by a single inspiratory capacity maneuver to total lung capacity, a five seconds breath hold and slow but maximal exhalation towards residual volume (17, 19, 20). Cross-contamination between subjects was prevented by a validated safety and disinfection protocol (see online supplement).

### Data-processing

Processing of raw eNose sensor signals included filtering, detrending, environmental correction and peak detection, which was performed automatically by the standard eNose software as published elsewhere (17, 19). For each measurement, two variables were selected for each sensor signal (sensor 1-7) that showed discriminative power in previous studies: (i) the highest sensor peak normalized to the most stable sensor (sensor 2) to minimize differences between sensor arrays and (ii) the ratio between the sensor peak and the breath hold (BH) point (17, 19).

### Statistical analysis

A priori sample size estimates indicated that approximately 30 patients per group is sufficient for building a diagnostic model (the training set) which can be validated in a new group of 30 individuals with SARS-CoV-2 infection (See online supplement) (13, 15, 16). SPSS (IBM, version 23) was used for statistical analysis. Descriptive statistics were summarized as either means with standard deviation (SD) or medians with interquartile range (IQR). Between-group comparisons were performed using two-sample unpaired t-tests, Mann–Whitney U tests or chi-squared tests.

Study participants in the *training* and *validation* set were included at the same test facility. By the creation of a new variable that selected a random sample (50:50) of cases, *training* and *validation* set were separated. In the *training* set, the normalized sensor peaks and peak/BH ratios were compared between groups using independent sample t-tests. The significance of the t-tests was determined using 1.000 iterations of the bootstrap. The variables that discriminated (p<0·05) between individuals with and without SARS-CoV-2 were used as input for linear discriminant analysis in the *training* set to calculate a discriminant function that best distinguished between the groups. The accuracy of this diagnostic model was defined as the percentage of correctly classified participants. Cross-validation of the linear discriminant analysis including the leave-one-out method was used to calculate the cross-validated value (CVV, %), keeping the selected predictor variables fixed. Subsequently, the cross-validated discriminant scores were used as input to construct receiver operating characteristics (ROC) curves with a 95% confidence interval (CI).

To test validity, the discriminant function obtained from the *training* set was examined in the *validation*, the *replication* and in the *asymptomatic set*, and used as input to construct ROC curves with a 95% confidence interval (CI). Discriminant scores were converted into a predicted percentage score for SARS-CoV-2 to aid interpretation. Finally, in the *training* set, we selected multiple cut-off points for the predicted SARS-CoV-2 percentage score to detect an infection with a sensitivity of approximately 100%, 97.5%, and 95%. Subsequently, the sensitivity, specificity, Positive Predicted Value (PPV) and Negative Predicted Value (NPV) for these cut-off points were calculated in the *validation* set. The discriminative ability of the ROC curves was summarized in area under the curves (AUC). See online supplement for more details. Additionally, the discriminant function obtained from the *training* set was validated in the external *replication* set including individuals recruited in a second test facility and in the *asymptomatic set*. Again the ROC curve with a 95% CI was constructed. For the *replication* set and in the asymptomatic set, one predefined cut-off point was used to calculate eNose test characteristics e.g. the sensitivity, specificity, PPV and NPV.

### Role of the funding source

The sponsor of the study had no role in study design, data collection, data analysis, data interpretation, or writing of the report. All authors had full access to all the data in the study and they all accept responsibility to submit for publication.

## Results

A total number of 4593 individuals participated in the study. Of these, 4510 were available for analysis (Figure 1). In the *training* and *validation* set, 1808 individuals were included in August 2020. Of these, 68 were SARS-CoV-2 positive (prevalence 3.8%). Of the SARS-CoV-2 positives, 65 had a positive molecular diagnostic test, either TMA or RT-PCR, at inclusion and three individuals had a positive RT-PCR test within 7 days after inclusion. All study participants survived at 30 days follow up.

**Figure 1.**
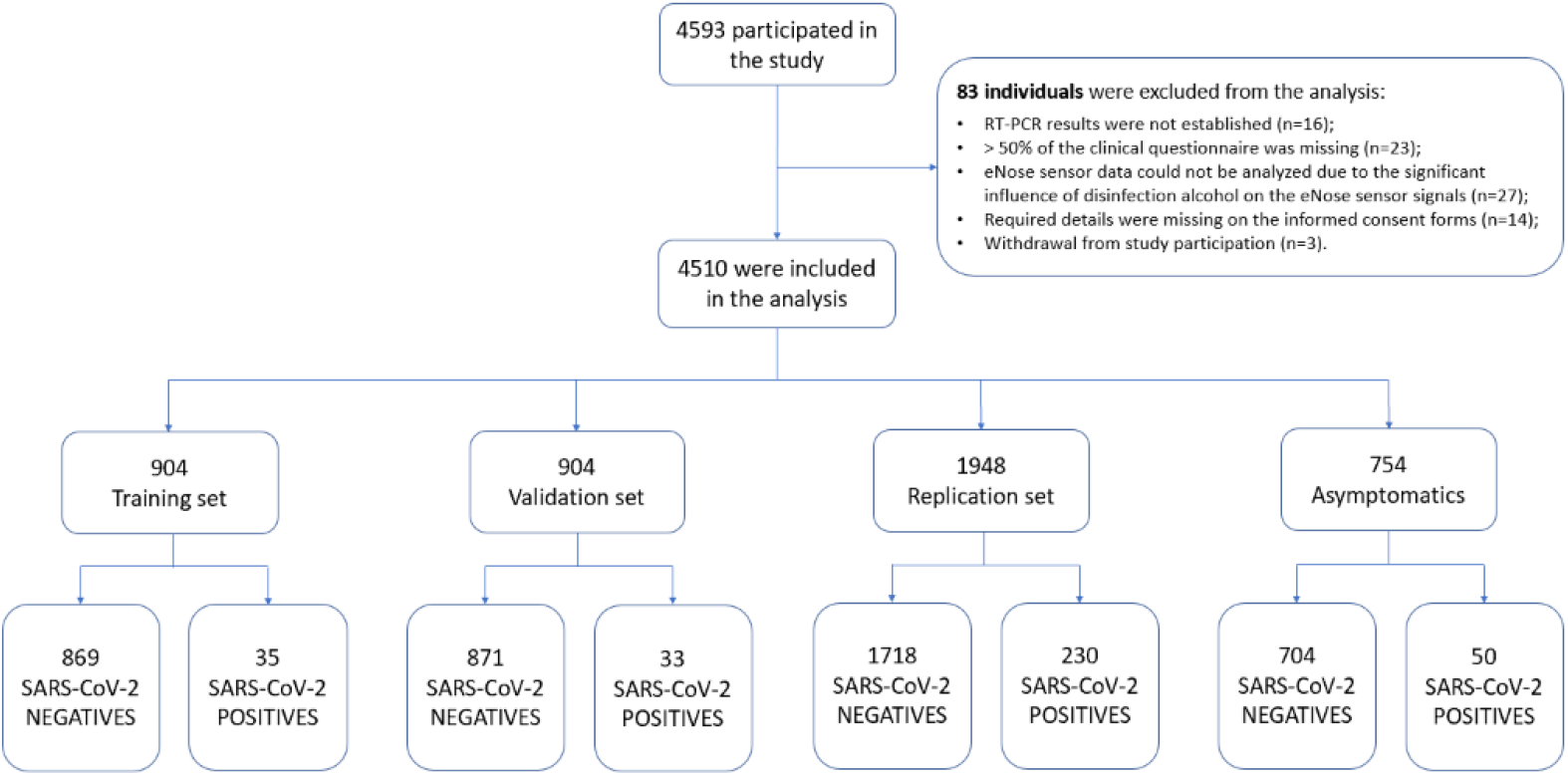
Participant enrolment

In the second test facility, 1948 individuals, of which 230 were SARS-CoV-2 positive, were included in the *replication* set in November 2020 (SARS-CoV-2 prevalence 11.8%). The *asymptomatic set* included 754 participants in both first and second test facility in December 2020. Fifty of them were SARS-CoV-2 positive (prevalence 6.6%) (Figure 1.).

Baseline characteristics are listed in Table 1. In the *training, validation* and *asymptomatic* set smoking status was significantly different (p<0.05) between the SARS-CoV-2 positive and negative group. In the *validation* set the SARS-CoV-2 positive group consisted in majority of males whereas the SARS-CoV-2 negative group consisted in majority of females.

**Table 1.** Baseline characteristics of individuals with and without a SARS-CoV-2 infection.

|  | Training set |  | Validation set |  | Replication set |  | Asymptomatic set |  |
| --- | --- | --- | --- | --- | --- | --- | --- | --- |
|  | SARS-CoV-2<br>POSITIVE<br>(N=35) | SARS-CoV-2<br>NEGATIVE<br>(N=869) | SARS-CoV-2<br>POSITIVE<br>(N=33) | SARS-CoV-2<br>NEGATIVE<br>(N=871) | SARS-CoV-2<br>POSITIVE<br>(N=230) | SARS-CoV-2<br>NEGATIVE<br>(N=1718) | SARS-CoV-2<br>POSITIVE<br>(N=50) | SARS-CoV-2<br>NEGATIVE<br>(N=704) |
| Male sex, no.(%) | 11 (31) | 383 (44) | 21 (64)<sup>a</sup> | 372 (43)<sup>a</sup> | 114 (50) | 764 (45) | 20 (40) | 332 (47) |
| Age (years), mean (SD) | 29.3 (9.6) | 33.4 (12.4) | 31.1 (12.3) | 33.3 (13.0) | 40.0 (14.6) | 39.0 (14.5) | 36.3 (13.7) | 34.1 (13.2) |
| BMI, mean (SD) | 22.0 (2.8) | 23.4 (5.5) | 24.2 (2.6) | 23.3 (3.5) | 25.5 (9.8) | 24.4 (6.9) | 23.6 (2.6) | 23.4 (3.5) |
| Smoking<br>(never/ex/current) | 19/5/11<sup>a</sup> | 462/189/218<sup>a</sup> | 14/8/11<sup>a</sup> | 449/182/240<sup>a</sup> | 128/66/36 | 940/472/306 | 25/13/11<sup>a</sup> | 427/144/132<sup>a</sup> |
| Pack years, mean (SD) | 0.8 (2.2) | 2.4 (7.4) | 1.0 (2.0) | 2.3 (6.5) | 4.1 (8.5) | 3.3 (8.4) | 2.1 (4.3) | 2.0 (7.6) |
| Asymptomatic, no. (%) | 3 (9) | 132 (15) | 5 (15) | 150 (17) | 35 (15) | 135 (8) | 50 (100) | 704 (100) |
| Fever, no.(%) | 2 (6) | 69 (8) | 1 (3) | 79 (9) | 10 (4) | 117 (7) | NA | NA |
| Comorbidity, no.(%) | 4 (11) | 136 (16) | 8 (24) | 132 (15) | 28 (12) | 188 (11) | 0 (0) | 46 (7) |
| VAS Dyspnea, mean (SD) | 11.9 (18.6) | 13.7 (18.9) | 15.4 (21.3) | 12.9 (17.6) | 12.0 (19.0) | 12.9 (20.7) | NA | NA |
| VAS Cough, mean (SD) | 15.4 (20.6) | 15.9 (19.3) | 19.9 (27.2) | 16.1 (19.6) | 18.2 (21.7) | 19.0 (23.0) | NA | NA |

<sup>a</sup> Significant difference between individuals with and without a SARS-CoV-2 infection ( $p < 0.05$ ). SD: Standard Deviation. BMI: Body Mass Index. VAS: Visual Analogue Scale (0-100mm). NA: Not Applicable

### Training set

The independent t-tests showed that the normalized sensor peaks of sensor 1 (p=0.001) and sensor 6 (p=0.001) and the peak/BH ratios sensor 3_BH (p=0.019), sensor 5_BH (p=0.001) and sensor 6_BH (p=0.001) were significantly different between individuals with and without a SARS-CoV-2 infection. Subsequent linear discriminant analysis yielded a cross-validated value of 84.3%. The AUC after internal cross-validation reached 0·948 (CI:0.929-0.967) (Figure 2A). No associations were found between eNose variables and Ct values.

**Figure 2.**
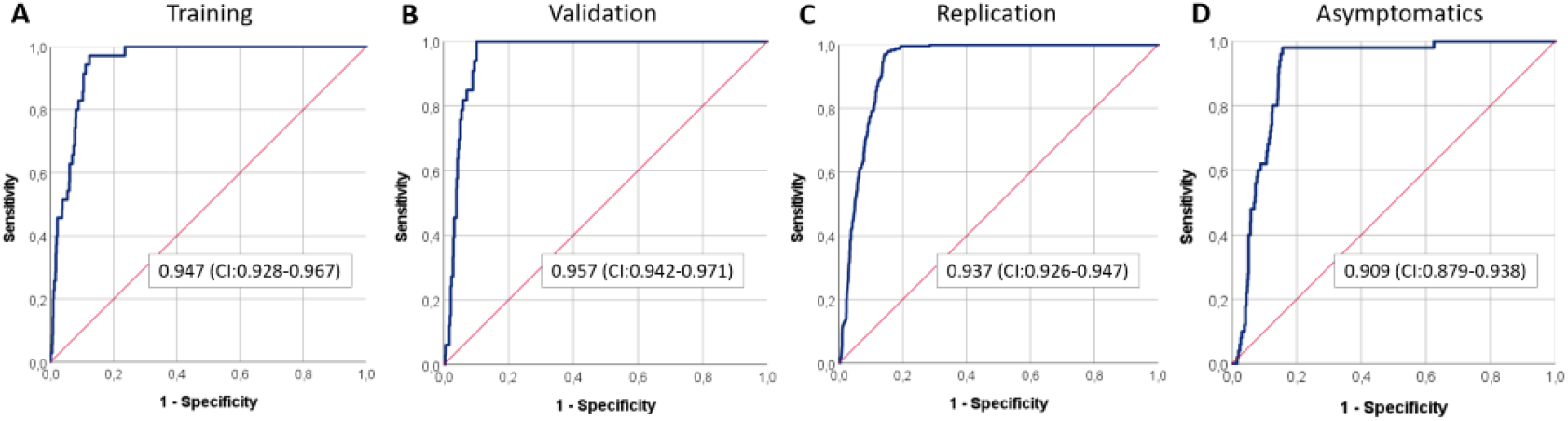
**A) Training set:** ROC curve with line of identity of the breath profile discriminant function (representing sensor 1, sensor 6, sensor 3_BH, sensor 5_BH and sensor 6_BH), predictive for the detection of a SARS-CoV-2 infection (area under the curve (AUC): 0.947). **B) Validation set:** ROC curve with line of identity of the breath profile discriminant function, obtained from the *training* set and predictive for the detection of a SARS-CoV-2 infection (AUC: 0.957). **C) Replication set:** ROC curve with line of identity of the breath profile discriminant function, obtained from the *training* set and predictive for the detection of a SARS-CoV-2 infection (AUC: 0.937). **C) Asymptomatic set:** ROC curve with line of identity of the breath profile discriminant function, obtained from the *training* set and predictive for the detection of a SARS-CoV-2 infection (AUC: 0.909).

### The diagnostic model

This mathematical model is the result of the linear discriminant analysis in the *training* set and is represented by the following equation. This equation represents an individual percentage score for a SARS-CoV-2 infection based on the eNose assessment. Labels S1 and S6 indicate the normalized sensor peak of sensor 1 and sensor 6. S3_BH_, S5_BH_ and S6_BH_ represent the ratio between the highest sensor peak and the BH point for sensors 3, 5 and 6.

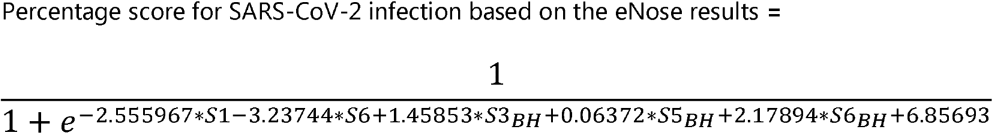

### Validation set

The ability to detect SARS-CoV-2 from exhaled breath using this model was confirmed in the *validation* set. Breath profiles of individuals with and without a SARS-CoV-2 infection were distinguished with a ROC-AUC of 0.957 (CI:0.942-0.971) (Figure 2B). The sensitivity, specificity, NPV and PPV of 3 cut-off values are listed in Table 2. To exclude false negatives, we selected a cut-off value of 0.30 for the percentage score for a SARS-CoV-2 infection based on the *training* set to obtain 100% sensitivity (Figure 3A). In the *validation* set, this cut-off value resulted in a sensitivity of 100%, a specificity of 78%, a NPV of 100% and a PPV of 15%. Based on the eNose results, 681 out of 904 individuals (75%) showed a percentage score of SARS-CoV-2 < 0.30. These 681 individuals were all tested negative for SARS-Cov-2 based on molecular amplification test results.

**Table 2.**
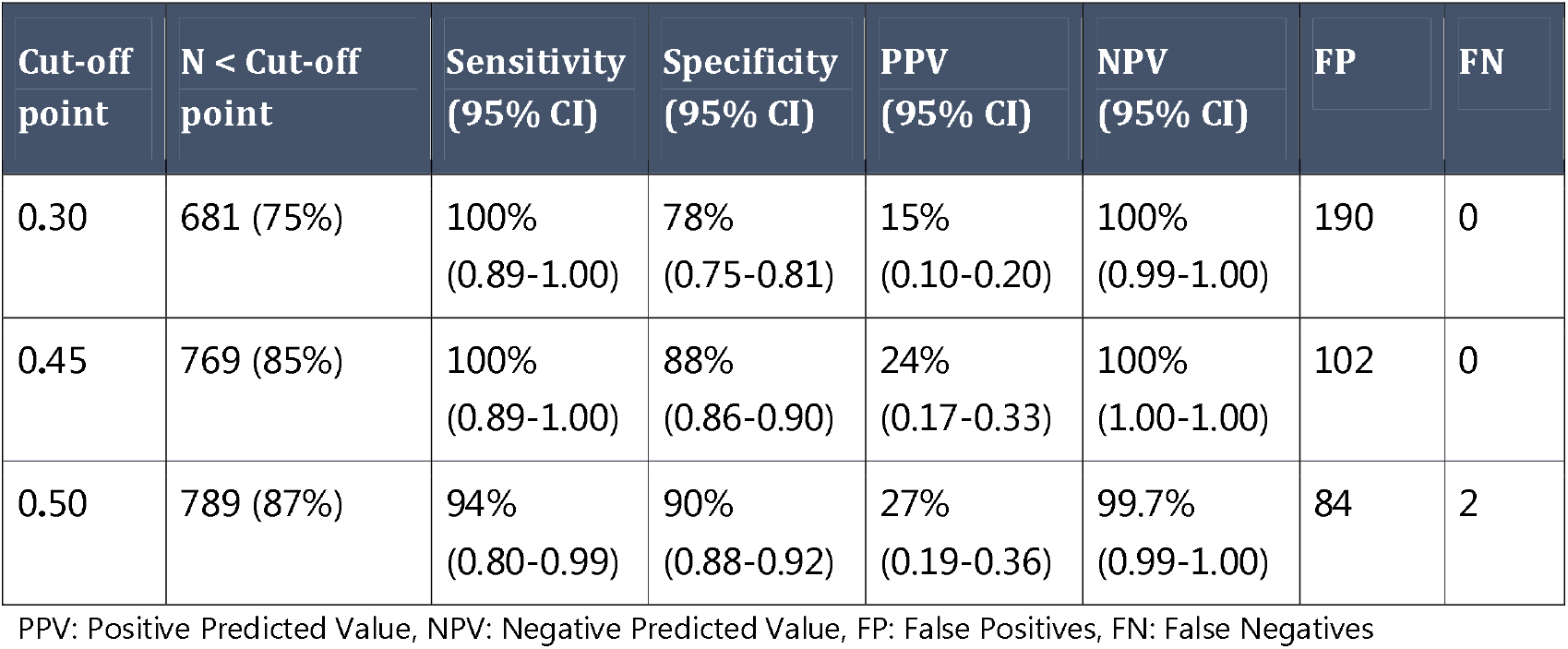
Test characteristics of the validation set. Multiple cut-off points for the detection of a SARS-CoV-2 infection that were selected from the training set and tested in the validation set.

| Cut-off point | N < Cut-off point | Sensitivity (95% CI) | Specificity (95% CI) | PPV (95% CI) | NPV (95% CI) | FP | FN |
| --- | --- | --- | --- | --- | --- | --- | --- |
| 0.30 | 681 (75%) | 100%<br>(0.89-1.00) | 78%<br>(0.75-0.81) | 15%<br>(0.10-0.20) | 100%<br>(0.99-1.00) | 190 | 0 |
| 0.45 | 769 (85%) | 100%<br>(0.89-1.00) | 88%<br>(0.86-0.90) | 24%<br>(0.17-0.33) | 100%<br>(1.00-1.00) | 102 | 0 |
| 0.50 | 789 (87%) | 94%<br>(0.80-0.99) | 90%<br>(0.88-0.92) | 27%<br>(0.19-0.36) | 99.7%<br>(0.99-1.00) | 84 | 2 |

PPV: Positive Predicted Value, NPV: Negative Predicted Value, FP: False Positives, FN: False Negatives

**Figure 3:**
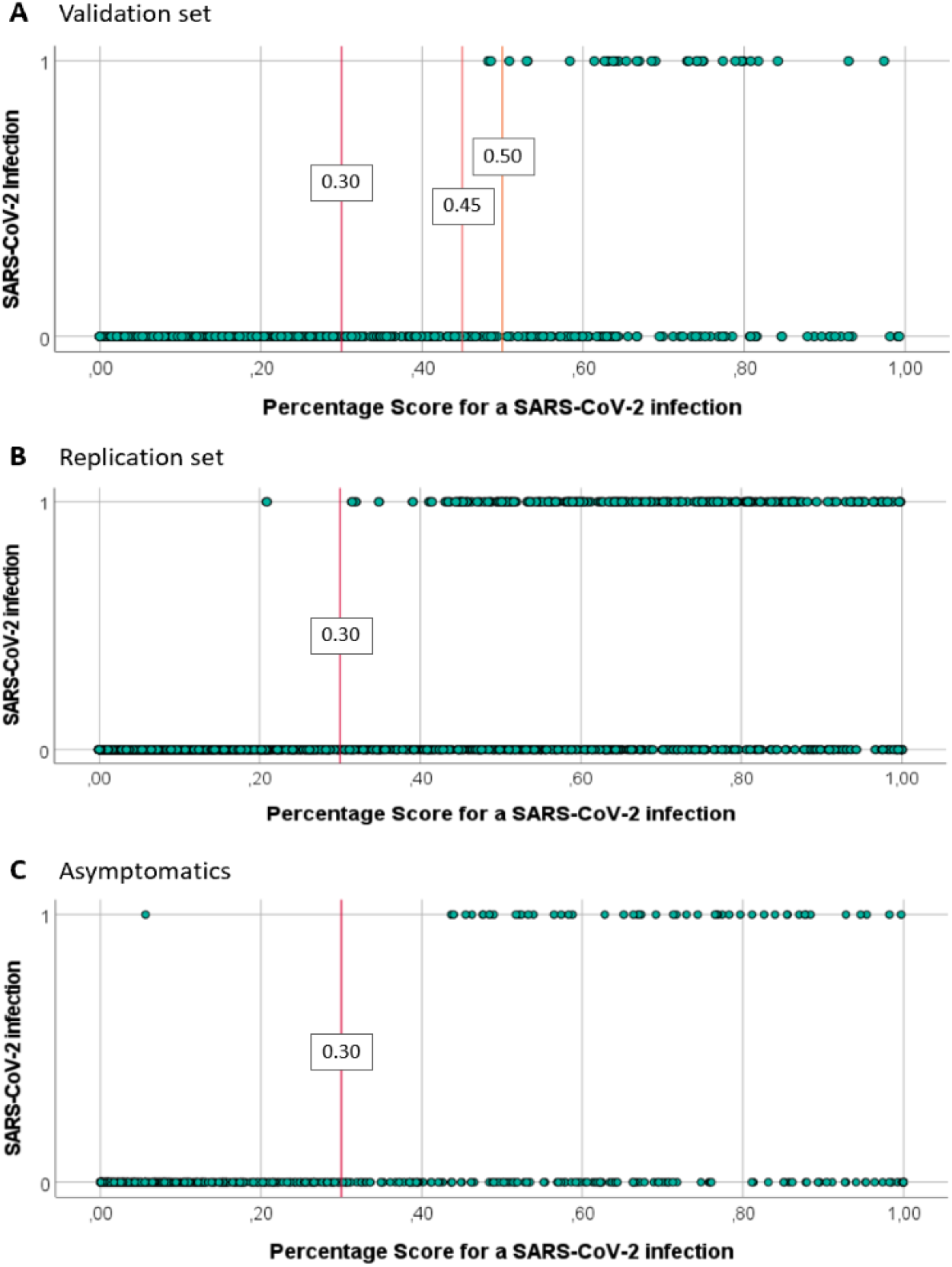
**A) Validation set:** Two-dimensional plot showing the percentage score for a SARS-CoV-2 infection obtained from the eNose results. A cut-off value of 0.30 was selected based on the training set in order to achieve 100% sensitivity for detection of SARS-CoV-2. The cut-off values of 0.45 and 0.50 were selected for comparison. **B) Replication set:** Two-dimensional plot showing the percentage score for a SARS-CoV-2 infection obtained from the eNose results. A cut-off value of 0.30 was selected based on the training set. **C) Asymptomatic set:** Two-dimensional plot showing the percentage score for a SARS-CoV-2 infection obtained from the eNose results. A cut-off value of 0.30 was selected based on the training set. 1: Individuals with a SARS-CoV-2 infection based on the classification definition, 0: Individuals without a SARS-CoV-2 infection.

### Replication set

External validation of the diagnostic model was performed in an independent *replication* set. Breath profiles of individuals with and without SARS-CoV-2 infection were distinguished with a ROC-AUC of 0.937 (CI:0.926-0.947) (Figure 2C). The selected cut-off value of 0.30 for the percentage score for a SARS-CoV-2 infection resulted in a sensitivity of 99.6%, a specificity of 79.8%, a NPV of 99.9% and a PPV of 39.9% (Table 3).

**Table 3.**
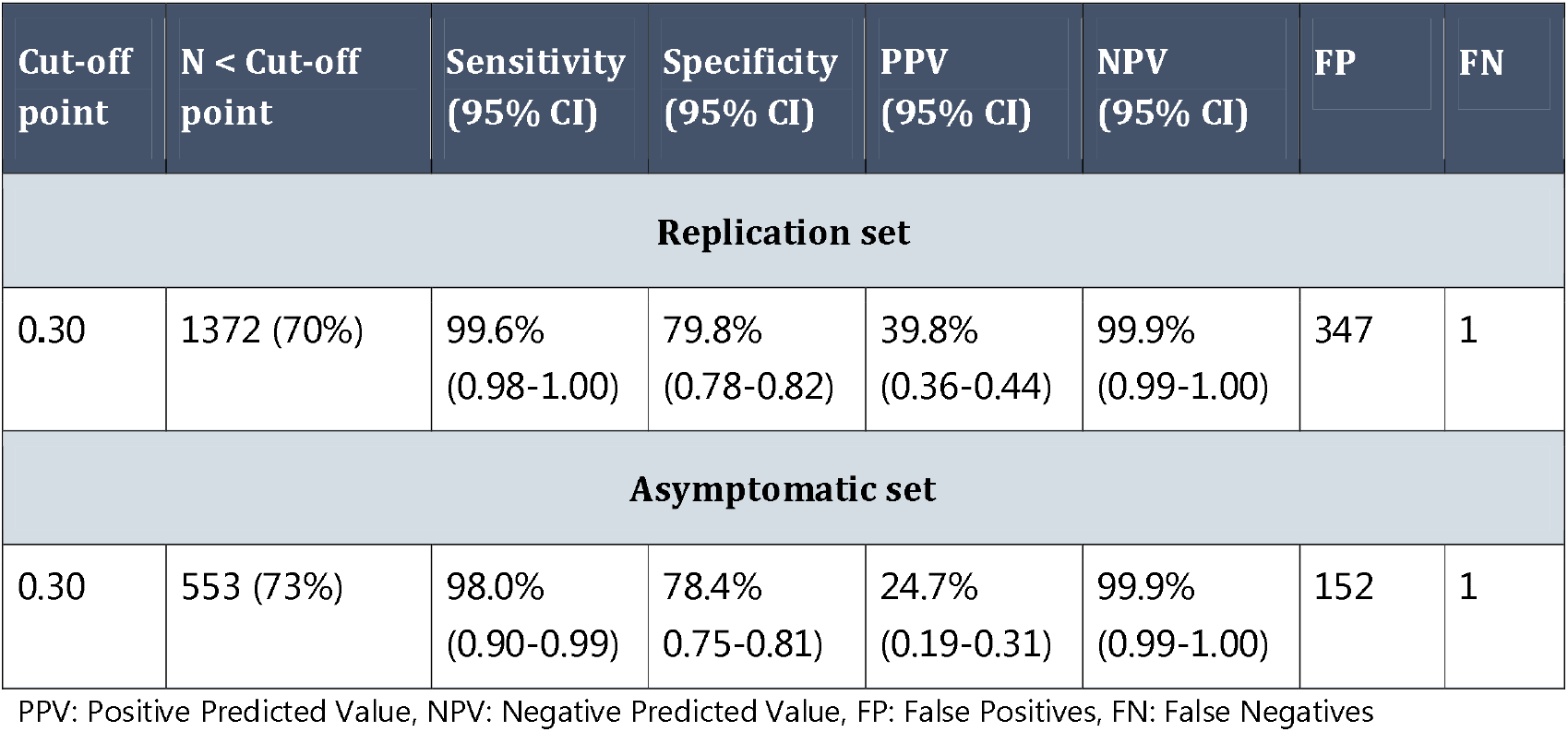
Test characteristics for the replication set and for the asymptomatic set. A cut-off value of 0.30 for the detection of a SARS-CoV-2 infection was selected based on the training set.

| Cut-off point | N < Cut-off point | Sensitivity (95% CI) | Specificity (95% CI) | PPV (95% CI) | NPV (95% CI) | FP | FN |
| --- | --- | --- | --- | --- | --- | --- | --- |
| **Replication set** |  |  |  |  |  |  |  |
| 0.30 | 1372 (70%) | 99.6%<br>(0.98-1.00) | 79.8%<br>(0.78-0.82) | 39.8%<br>(0.36-0.44) | 99.9%<br>(0.99-1.00) | 347 | 1 |
| **Asymptomatic set** |  |  |  |  |  |  |  |
| 0.30 | 553 (73%) | 98.0%<br>(0.90-0.99) | 78.4%<br>(0.75-0.81) | 24.7%<br>(0.19-0.31) | 99.9%<br>(0.99-1.00) | 152 | 1 |

PPV: Positive Predicted Value, NPV: Negative Predicted Value, FP: False Positives, FN: False Negatives

### Asymptomatic set

Breath profiles of asymptomatic individuals with and without a SARS-CoV-2 infection were distinguished with a ROC-AUC of 0.909 (CI: 0.879-0.938) (Figure 2D). The selected cut-off value of 0.30 for the percentage score for a SARS-CoV-2 infection resulted in a sensitivity of 98.0%, a specificity of 78.4%, a NPV of 99.9% and a PPV of 24.7% (Table 3).

## Discussion

This was a prospective real-world study that evaluated the performance of a cloud-connected eNose to detect or rule out a SARS-CoV-2 infection. The results show that exhaled breath analysis by eNose allows discrimination between individuals with and without SARS-CoV-2 and can reliably rule out an infection in 70-75% of those who visit the public health test facilities. These findings were confirmed in an *validation* set and externally validated in an independent *replication* and *asymptomatic* set.

To our knowledge this is the first population-based, large scale study examining the capacity of an eNose system to detect SARS-CoV-2 infection. The potential ability of exhaled breath analysis by sensor-based technologies for COVID-19 diagnosis has been demonstrated by three pilot studies including a pre-diagnosed population (1, 2, 4). The obtained signals during these pilots provided adequate discrimination between patients with and without COVID-19 with ROC-AUCs ranging between 0.74 and 0.94. Our study extends this work to a large real-world setting by including a population of symptomatic and asymptomatic individuals, which represent the population in public health care testing programs worldwide.

Presence of VOC signatures of viral infections have been demonstrated in both *in-vitro* and *in-vivo* studies, already before the pandemic caused by SARS-CoV-2 (21-25). VOC formation during an infection is the result of a multi-step and highly complex interaction between virus and host response (15). Notably, exhaled VOC signatures seem to be virus-specific (21, 22, 26). This could be partly due to the fact that various types of viruses use different receptors for cell entry; for example, SARS-CoV-2 binds the angiotensin-converting enzyme 2 (ACE2) receptor (27), whereas seasonal influenza viruses mainly use sialic acids receptors (22). This leads to distinct downstream pathways within the infected cells, potentially followed by formation of virus-specific VOCs even at early stages of the disease (7, 12). Although initial symptoms of SARS-CoV-2 infection may resemble other viral respiratory infections, COVID-19 is complicated by extra-pulmonary manifestations such as gastro-intestinal symptoms, acute kidney injury, and thrombotic complications (28), which are not common in other respiratory viral infections. Finally, SARS-CoV-2 demonstrates the ability to evade innate immunity and induce cytokine hyper production (29) uncommon for seasonal respiratory viruses. All these interactions may result in different VOC patterns distinguishing SARS-CoV-2 from other viral respiratory pathogens. This underlines the potential of eNose technology as a diagnostic tool in future outbreaks of infectious diseases.

Smoking is described to influence the composition of exhaled VOCs and therefore, is considered as a potentially confounding factor in exhaled breath analysis (30). In both the *training, validation* and *asymptomatic* set of this study smoking status was significantly different between the SARS-CoV-2 positive and negative group. When predicting eNose variables based on smoking status, no significant regression equations were found (see online supplement). Therefore, it is unlikely that smoking had an influence on our results.

The present findings were obtained by integrative analysis of cross-reactive sensors that are not selective with regard to individual VOCs (14). COVID-19 specific VOC signatures in exhaled breath have been identified with approaches other than sensor-based technologies. Interestingly, despite the broad differences between study designs, used technologies, and exhaled breath measurements, all seven available studies including ours demonstrate adequate accuracies for detecting SARS-CoV-2 infection (1, 2, 4, 26, 31, 32). Three recent studies using chemical analytical technologies have found several aldehydes (e.g. butyraldehyde, ethanal, methylpent-2-enal, nonanal), esters (ethyl butanoate), ketones (e.g. acetone), alcohols (e.g. isopropanol, methanol) and an alkadiene (2,4-octadiene) to differentiate between those with and without COVID-19 (4, 31, 32). Furthermore, in a small study using a multi-capillary column coupled ion mobility spectrometry (MCC-IMS), Steppert *et al*. have recently demonstrated that breath profiles of those infected with SARS-Cov-2 are significantly different from those with Influenza A infection (26).

Among different technologies, eNoses are specifically designed to capture a complete mixture of VOCs, which takes advantage of information all along the VOC spectrum. This allows for composite biomarker discovery rather than measuring individual molecules that is needed when aiming to identify the underlying molecular networks. The eNose used in the current study is coupled to a cloud-solution, which applies machine learning algorithms to search for patterns in the obtained sensor signals. Pattern recognition allows for probabilistic analyses, which result in the present ROC-curves that can be used for clinical diagnostics. Notably, depending on the investigated disease, a different cut-off value and thereby, different sensitivity and specificity can be selected for decision making. This is where the advantage of a percentage score for SARS-CoV-2 infection, compared to a binary (yes/no) test outcome, becomes apparent. In this study in subjects tested at a public health care facility, a cut-off value of 0.30 selected from the *training* set resulted in 100% sensitivity and 78% specificity in the *validation* set, 99.6% sensitivity and 83.9% specificity in the *replication* set and 98.0% sensitivity and 82.2% specificity in the *asymptomatic* set. Both in the *replication* set and in the *asymptomatic* set, one false negative eNose result was present. Both cases had low viral loads (CT value 42.3 and 31.7 respectively), and therefore were most likely unable to infect others. The high sensitivity is vital to minimize the risk of further spread of the virus. This approach further qualifies the eNose as a screening test before more specific secondary molecular amplification or antigen tests. The results of our study demonstrate that in a scenario where eNose is used as a screening test, swab sampling for molecular amplification tests could be avoided with a safe margin in 70-75% of those who visit the test facilities. This could subsequently reduce workload and burden on the laboratories substantially and increase residual testing capacity, thereby significantly changing the SARS-CoV-2 test procedure in public health care setting.

An important strength of our study is the unselected population-based setting in which individuals were included during day-to-day visits to the test facility without prior diagnosis of COVID-19, which enhances the generalizability of our results. Second, the combination of a *validation* set, an independent *replication* set and a set with asymptomatic individuals that confirmed the results of the *training* set strengthen the observed accuracies. And finally, a cloud-connected eNose was used that is both technically and clinically validated (17-20). Connection of multiple eNoses to a cloud solution integrated with artificial intelligence and a large database makes it suitable for implementation at the point-of-care as the results can be made available within seconds.

One of the limitations of the study could be that a SARS-CoV-2 infection was merely based on the results of molecular amplification tests. This could explain some of the false positives found by the eNose as there is concern about false negative RT-PCR results for detecting SARS-CoV-2 (33). In our *validation* set, including follow-up (≤ 7 days) TMA or RT-PCR results to our reference standard, we were able to add some initial false negatives to the positive group, only partially mitigating the suboptimal sensitivity of the initial SARS-CoV-2 RNA detection test. During this study in the Netherlands, very few other commonly known viruses were circulating, besides rhinovirus. Due to the real-life setting and timing of the study, we cannot yet provide an answer about the accuracy of the eNose for distinguishing SARS-CoV-2 from other respiratory viruses. Given that these viruses harbor unique viral proteins, different receptor preferences, and distinct interactions with hosts on a cellular level, specific VOC patterns could possibly be identified for various viral respiratory pathogens. Finally, the present population mostly comprised young ambulant adults that visit public test facilities but that may not be representative of older and in-hospital populations.

In conclusion, eNose technology can distinguish between individuals with and without a SARS-CoV-2 infection with high accuracy. Exhaled breath analysis by eNose represents a quick and non-invasive method to reliably rule out a SARS-CoV-2 infection in public health test facilities. Using the eNose as a screening test can reduce the number of throat and nasopharyngeal swabs and the required SARS-CoV-2 RNA detection tests, which in turn can reduce the burden on individuals, economy and healthcare.

## Supporting information

Online supplement eNose Covid-19 study

STARD 2015 checklist

## Data Availability

Data sharing
The study methods, statistical analysis plan, and detailed analysis are available in the main Article and the online supplement. Deidentified participant data and the protocol are available on reasonable request.

## Contribution statement

RV, RMV, JJG, LGV, ML, PJS, JCCMV and GHG designed the study; RMV, SM, DRV, SB, BC, JCCMV and GHG did the data collection; RV, JJG, LGV, PJS, JCCMV and GHG did the data analysis; RV, JJG and GHG verified the underlying data; RV, RMV, JJG, SB, JJMG, LGV, PJS, JCCMV and GHG did the data interpretation; RV, RMV, DRV, PJS, JCCMV and GHG wrote the report; All the authors critically revised the manuscript, vouch for the accuracy and completeness of the data reported and all the authors made the decision to submit the manuscript for publication. All authors read and approved the final manuscript.

## Declaration of interests

RV receives personal fees and has a substantial interest in the start-up company Breathomix BV. NF receives personal fees from the start-up company Breathomix BV. PJS is scientific adviser and has an officially non-substantial interest in the start-up company Breathomix BV. All other authors have no conflicts of interest.

## Data sharing

The study methods, statistical analysis plan, and detailed analysis are available in the main Article and the *online supplement*. Deidentified participant data and the protocol are available on reasonable request.

## Acknowledgements

This study was funded by Ministry of Health, Welfare and Sport of the Netherlands. The sponsor of the study had no role in study design, data collection, data analysis, data interpretation, or writing of the report. The authors wish to acknowledge Veerle Roukens, Sam van Hugten and Fleur Storm van Leeuwen at the Leiden University Medical Center for retrieving the follow up data and including individuals in the replication cohort and asymptomatic cohort.

## Notes

### Clinical Trial

Prospective observational cohort

### Author Declarations

The study protocol was approved by the Medical Ethics Committee of Leiden The Hague Delft (P20.033).

## References

1. Shan B, Broza YY, Li W, Wang Y, Wu S, Liu Z, et al. Multiplexed Nanomaterial-Based Sensor Array for Detection of COVID-19 in Exhaled Breath. ACS Nano. 2020;14(9):12125–32.

2. Wintjens A, Hintzen KFH, Engelen SME, Lubbers T, Savelkoul PHM, Wesseling G, et al. Applying the electronic nose for pre-operative SARS-CoV-2 screening. Surg Endosc. 2020:1–8.

3. Berna AZ, Akaho EH, Harris RM, Congdon M, Korn E, Neher S, et al. Breath biomarkers of pediatric SARS-CoV-2 infection: a pilot study. medRxiv. 2020.

4. Grassin-Delyle S, Roquencourt C, Moine P, Saffroy G, Carn S, Heming N, et al. Metabolomics of exhaled breath in critically ill COVID-19 patients: A pilot study. EBioMedicine. 2021;63:103154.

5. European Centre for Disease Prevention and Control. COVID-19 testing strategies and objectives. 15 September 2020. ECDC: Stockholm; 2020.

6. Gorzalski AJ, Tian H, Laverdure C, Morzunov S, Verma SC, VanHooser S, et al. High-Throughput Transcription-mediated amplification on the Hologic Panther is a highly sensitive method of detection for SARS-CoV-2. J Clin Virol. 2020;129:104501-.

7. Russo A MC, Starace M, Astorri R, Calò F, Coppola N. Current Status of Laboratory Diagnosis for COVID-19: A Narrative Review. Infect Drug Resist. 2020(13):2657–65

8. WHO. Laboratory testing for coronavirus disease (COVID-19) in suspected human cases. [Available from: https://www.who.int/publications-detail/laboratory-testing-for-2019-novel-coronavirus-in-suspected-human-cases-20200117.

9. Feng W, Newbigging AM, L. C, Pang B, Peng H, Cao Y, et al. Molecular Diagnosis of COVID-19: Challenges and Research Needs. Anal Chem. 2020;92(15):10196–209.

10. Brooks SK, Webster RK, Smith LE, Woodland L, Wessely S, Greenberg N, et al. The psychological impact of quarantine and how to reduce it: rapid review of the evidence. Lancet. 2020;395(10227):912–20.

11. Tromberg BJ, Schwetz TA, Pérez-Stable EJ, Hodes RJ, Woychik RP, Bright RA, et al. Rapid Scaling Up of Covid-19 Diagnostic Testing in the United States — The NIH RADx Initiative. New England Journal of Medicine. 2020;383(11):1071–7.

12. Gould O, Ratcliffe N, Król E, de Lacy Costello B. Breath analysis for detection of viral infection, the current position of the field. J Breath Res. 2020;14(4):041001.

13. de Vries R, Sterk PJ. eNose breathprints as composite biomarker for real-time phenotyping of complex respiratory diseases. J Allergy Clin Immunol. 2020.

14. Wilson AD. Advances in electronic-nose technologies for the detection of volatile biomarker metabolites in the human breath. Metabolites. 2015;5(1):140–63.

15. Lamote K, Janssens E, Schillebeeckx E, Lapperre TS, De Winter BY, van Meerbeeck JP. The scent of COVID-19: viral (semi-)volatiles as fast diagnostic biomarkers? J Breath Res. 2020;14(4):042001.

16. Farraia MV, Cavaleiro Rufo J, Paciência I, Mendes F, Delgado L, Moreira A. The electronic nose technology in clinical diagnosis: A systematic review. Porto Biomed J. 2019;4(4):e42.

17. de Vries R, Dagelet YWF, Spoor P, Snoey E, Jak PMC, Brinkman P, et al. Clinical and inflammatory phenotyping by breathomics in chronic airway diseases irrespective of the diagnostic label. Eur Respir J. 2018;51(1).

18. Moor CC, Oppenheimer JC, Nakshbandi G, Aerts JGJV, Brinkman P, Maitland – van der Zee A-H, et al. Exhaled breath analysis by use of eNose technology: a novel diagnostic tool for interstitial lung disease. European Respiratory Journal. 2020:2002042.

19. de Vries R, Muller M, van der Noort V, Theelen W, Schouten RD, Hummelink K, et al. Prediction of response to anti-PD-1 therapy in patients with non-small-cell lung cancer by electronic nose analysis of exhaled breath. Ann Oncol. 2019;30(10):1660–6.

20. de Vries R, Brinkman P, van der Schee MP, Fens N, Dijkers E, Bootsma SK, et al. Integration of electronic nose technology with spirometry: validation of a new approach for exhaled breath analysis. J Breath Res. 2015;9(4):046001.

21. Schivo M, Aksenov AA, Linderholm AL, McCartney MM, Simmons J, Harper RW, et al. Volatile emanations from in vitro airway cells infected with human rhinovirus. J Breath Res. 2014;8(3):037110.

22. Aksenov AA, Sandrock CE, Zhao W, Sankaran S, Schivo M, Harper R, et al. Cellular scent of influenza virus infection. Chembiochem. 2014;15(7):1040–8.

23. Purcaro G, Rees CA, Melvin JA, Bomberger JM, Hill JE. Volatile fingerprinting of Pseudomonas aeruginosa and respiratory syncytial virus infection in an in vitro cystic fibrosis co-infection model. J Breath Res. 2018;12(4):046001.

24. van Geffen WH, Bruins M, Kerstjens HA. Diagnosing viral and bacterial respiratory infections in acute COPD exacerbations by an electronic nose: a pilot study. J Breath Res. 2016;10(3):036001.

25. van der Schee MP, Hashimoto S, Schuurman AC, van Driel JS, Adriaens N, van Amelsfoort RM, et al. Altered exhaled biomarker profiles in children during and after rhinovirus-induced wheeze. Eur Respir J. 2015;45(2):440–8.

26. Steppert C, Steppert I, Sterlacci W, Bollinger T. Rapid detection of SARS-CoV-2 infection by multicapillary column coupled ion mobility spectrometry (MCC-IMS) of breath. A proof of concept study. medRxiv. 2020:2020.06.30.20143347.

27. Zhou P, Yang XL, Wang XG, Hu B, Zhang L, Zhang W, et al. A pneumonia outbreak associated with a new coronavirus of probable bat origin. Nature. 2020;579(7798):270–3.

28. Gupta A, Madhavan MV, Sehgal K, Nair N, Mahajan S, Sehrawat TS, et al. Extrapulmonary manifestations of COVID-19. Nature Medicine. 2020;26(7):1017–32.

29. Perico L, Benigni A, Casiraghi F, Ng LFP, Renia L, Remuzzi G. Immunity, endothelial injury and complement-induced coagulopathy in COVID-19. Nature Reviews Nephrology. 2020.

30. Dragonieri S, Pennazza G, Carratu P, Resta O. Electronic Nose Technology in Respiratory Diseases. Lung. 2017;195(2):157–65.

31. Ruszkiewicz D, Sanders D, O’Brien R, Hempel F, Reed M, Riepe A, et al. Diagnosis of COVID-19 by Analysis of Breath with Gas Chromatography-Ion Mobility Spectrometry - A Feasibility Study. SSRN Electronic Journal. 2020.

32. Chen H, Qi X, Ma J, Zhang C, Feng H, Yao M. Breath-borne VOC Biomarkers for COVID-19. medRxiv. 2020:2020.06.21.20136523.

33. Woloshin S, Patel N, Kesselheim AS. False Negative Tests for SARS-CoV-2 Infection — Challenges and Implications. New England Journal of Medicine. 2020;383(6):e38.

