## Supplementary material for "Ruling out SARS-CoV-2 infection using exhaled breath analysis by electronic nose in a public health setting": Online supplement eNose Covid-19 study

Table of Contents


- Measurement set up Page 2
- To ensure minimum risk of cross-contamination, the following safety measures were taken Page 3
- Follow-up data training and validation set Page 4
- Sample size estimation Page 5
- Bivariate regression analysis - Smoking status as determinant on eNose data Page 5
- Mathematical derivation of the percentage score for SARS-CoV-2 infection Page 6
- References Page 8

### Measurement set up

Exhaled breath analysis was performed using a cloud-connected eNose (SpiroNose), to detect the complete mixture of volatile organic compounds (VOCs), a Gateway to securely send data to an online server and an online analysis platform for real-time and automated analysis (Figure S1) (1,2). The SpiroNose includes 7 different metal oxide semiconductor (MOS) sensors and each sensor is present in duplicate on both the inside and outside of the eNose. The sensors on the inside of the eNose generate a breath profile, reflecting the collective VOC composition in exhaled breath. The sensors on the outside measure the complete mixture of ambient VOCs. The analysis platform has been developed according to the following standards: ISO 27001 (Information Security) and NEN 7510 (Information Security in healthcare).

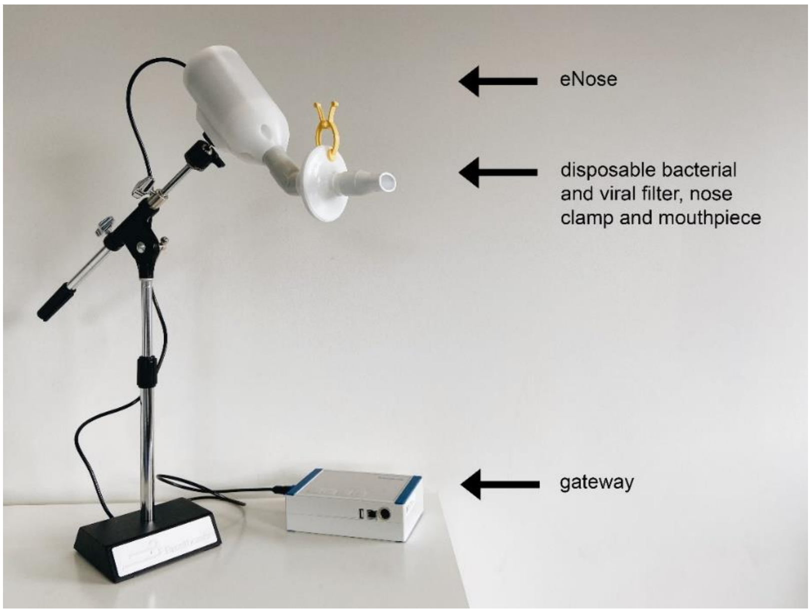


**Figure S1.** Measurement setup of the SpiroNose and its connection to the Gateway. Sensor data are securely transferred to the online analysis platform through the Gateway. The setup also includes a mouthpiece, nose clamp and viral/bacterial filter.

To ensure minimum risk of cross-contamination, the following safety measures were taken:

- All breath measurements were performed with a mouthpiece, nose clip and bacteria/virus filter (Pulmosafe) provided by Lemon Medical® (https://lemonmed.com/products/pulmosafe).
- This bacteria/virus filter consists of type separet 2402 of Freudenberg Filtration Technologies SE & Co, which has the highest protection grade (FFP3) and conforms to EU norm EN 149. It therefore offers protection against SARS-CoV-2 and blocks aerosols. After each measurement, the filter, mouthpiece and nose clamp were disposed.
- It has been shown that coronavirus can be effectively inactivated by surface disinfection using Hydrogen peroxide (0.5%) within 1 minute (3). Therefore, after every breath measurement, the outside and openings of the eNose were cleaned using a damp microfiber cloth and disinfected using 1.0 g Hydrogen peroxide (Incidin Oxywipes). This cleaning method was approved by the LUMC Infection Prevention department.
- The inside of the eNose is always heated (temperature: 50-60$^{o}$C) which consequently prevents formation of condensate that could potentially contain microbes. The sensors itself are heated up to 400 $^{o}$C. The virus will not survive these temperatures. In addition, the high temperature within the eNose results in a chimney effect that prevents ingress of dust and other particles into the device.
- All personnel using the eNose were trained in order to follow all the above safety measures and comply with the hygiene protocol that was created by the research team and approved by the LUMC Infection Prevention.

### Follow-up data training and validation set

From the 1808 subjects that were included in the analysis follow-up data of 1607 (89%) individuals were available. Hundred seventy-two individuals were lost to follow-up and 29 did not want to participate in the follow-up. From individuals that could not be reached via telephone 30 days after inclusion, data regarding 30-day mortality was obtained via the Dutch Central Bureau for Statistics (CBS) (4). There were in total 9 participants of whom data could not be retrieved, which was due to either missing or incorrect phone numbers, missing, incorrect or foreign citizen service numbers, or both.

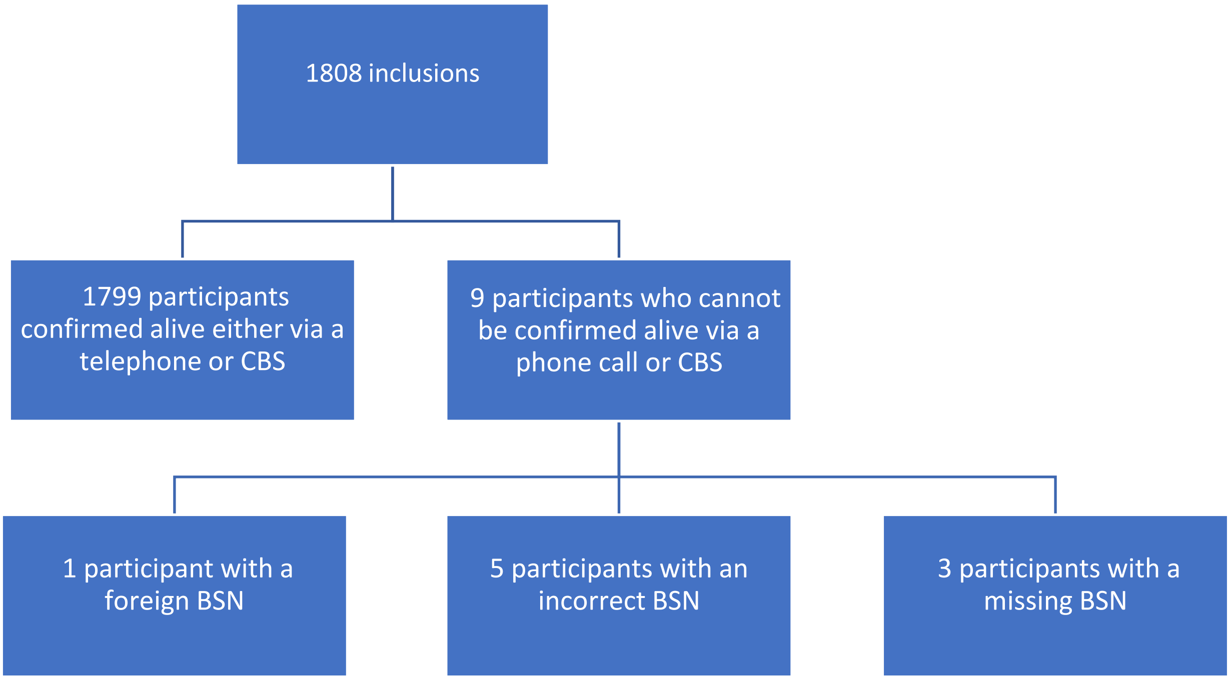


**Figure S2.** Mortality check

### Sample size estimation

As this is a first study, we are unable to perform a calculated sample size analysis. From a previous studies using the SpiroNose (1, 2) we have learned that approximately 20-30 patients per group is sufficient for building a diagnostic model that can be tested in a new group of individuals. In this study, we aimed to include at least 30 individuals with a SARS-CoV-2 infection in the *training* set and compare their breath profiles to 30 individuals without a SARS-CoV-2 infection. As this study was performed in a population of predominantly mild symptomatic individuals without prior SARS-CoV-2 infection, we could not tell which study participant was SARS-CoV-2 positive or negative at inclusion. Therefore, the group of SARS-CoV-2 negative participants was expected to end up much larger than needed for our analysis (e.g. the incidence rate for a SARS-CoV-2 infection at inclusion was ~4%, therefore we needed to include 25 individuals in order to include one individual with a SARS-CoV-2 infection). For the *validation* set, we estimated that the same number of individuals is sufficient: at least 30 individuals with and 30 individuals without a SARS-CoV-2 infection. The sample size of the *replication* set and the *asymptomatic* set was estimated based on the sensitivity obtained in the *training* set and the acceptable margin of uncertainty. For the obtained sensitivity of 0.99, a study population including 230 individuals with SARS-CoV-2 infection would result in a margin of uncertainty of 1.3% for the sensitivity and a study population of 50 SARS-CoV-2 positives would result in a margin of uncertainty of 2.4%.

### Bivariate regression analysis - Smoking status as determinant on eNose data

A simple linear regression was calculated in the *training* set to predict eNose variables (Sensor 1, Sensor 6, Sensor 3_BH, Sensor 5_BH and Sensor 6_BH) based on smoking status. No significant regression equations were found, meaning that smoking status did not contribute significantly to the prediction of the eNose variables.

### Mathematical derivation of the percentage score for SARS-CoV-2 infection

A percentage score for SARS-CoV-2 infection was calculated from the linear discriminant score in the training set and used to aid interpretation of results in the validation set, replication set and in the *asymptomatic* set. Since the percentage score is an increasing function of the discriminant score, the two measures contain exactly the same diagnostic information, and consequently they have identical ROC curves. However, from a clinical perspective the percentage scores are easier to interpret. The mathematical derivation of the equation for the percentage scores (2) consists of the following steps. These steps are standard for linear discriminant analysis (5).

#### 1. Discriminant score

After variable selection, linear discriminant analysis yields for each individual a discriminant score *D*:

D (individual) = $1.444 * S1 + 1.829 * S6 - 0.824 * S3_{BH} - 0.036 * S5_{BH} - 1.231 * S6_{BH} - 2.876$

The labels *S1* and *S6* represent the highest sensor peak of sensor 1 and sensor 6, respectively, both normalized to the most stable sensor, sensor 2. $S3_{BH}$, $S5_{BH}$ and $S6_{BH}$ indicate the ratio between the highest sensor peak and the breath hold (BH) point for sensor 3, 5 and sensor 6. The expression for *D* represents the most discriminative function between individuals with and without SARS-CoV-2 infection. In the training set the SARS-CoV-2 positive group has a mean D-score of 1.701817 and the SARS-CoV-2 negative group has a mean D-score of -0.068243.

#### 2. Percentage score for a SARS-CoV-2 infection

Percentage score for a SARS-CoV-2 infection (individual) = $\frac{1}{1+ e^{-\mathrm{L1}\ (individual)}}$

The percentage score for a SARS-CoV-2 infection, is computed from the *logit* of SARS-CoV-2 positives (*L1*) by the aforementioned equation. Conversely the *logit* can be computed from the training set probability; L1 = ln(Probability of SARS-CoV-2 positives/Probability of SARS-CoV-2 negatives), where ln is the natural logarithm. The relation between *L1* for each individual and their value of *D* is explained in the next step.

#### 3. Logit score

We adjusted the discriminant scores for the prior score using the following formula (5).

L1 (individual) = $d * D(individual) - d * s/2 + prior\ logit$

*D* represents the discriminant score of the study participant as calculated in the first step. Labels *d* and *s* represent the difference and sum of the means of the D-scores in both groups in the training set (SARS-CoV-2 positive: 1.701817 and SARS-CoV-2 negative: -0.068243). In this study d=1.77006 and s=1.633574. The *prior logit* symbolizes the logit of the probability of membership in the SARS-CoV-2 positive group before calculation. In the training set 35 SARS-CoV-2 positives and 869 SARS-CoV-2 negatives were included, hence giving a prior probability of a SARS-CoV-2 infection of 35/904 and a corresponding prior logit of ln(35/869)=-3.212.

L1 (individual) = $1.77006 * D(individual) - 1.76623$

$= 2.555967 * S1 + 3.23744 * S6 - 1.45853 * S3_{BH} - 0.06372 * S5_{BH} - 2.17894 * S6_{BH} - 6.85693$

The rationale for this is that the prior odds of having a SARS-CoV-2 infection are multiplied by a likelihood ratio to obtain the posterior odds of SARS-CoV-2 after taking into account the D-score. The likelihood ratio leading to this number is defined as: the probability density of SARS-CoV-2 positives given the value of the individual D score divided by the probability density of SARS-CoV-2 negatives. These probabilities are modeled by a normal distribution on the D scale with means at the group means 1.701817 and -0.068243 and a common standard deviation of 1, resulting from the normalization of the D-score. Describing what this means on the logit scale resulted in the above formula with *d* and *s*.

### 4. Percentage score for a SARS-CoV-2 infection (eNose)

Percentage score for a SARS-CoV-2 infection based on the eNose results **=**

$$= \frac{1}{1 + e^{-2.555967*S1 - 3.23744*S6 + 1.45853*S3_{BH} + 0.06372*S5_{BH} + 2.17894*S6_{BH} + 6.85693}}$$

With this equation, the percentage score for SARS-CoV-2 positives can be calculated for new participants. While the intent of the percentage score is to obtain a measure that approximates the estimated probability of having a SARS-CoV-2 infection, in this study we assessed only the discriminatory power of the scores, and not yet the calibration of the percentage score as a probability. We do not claim a probabilistic interpretation of the percentage score at this stage.

### References


1. de Vries R, Dagelet YWF, Spoor P, Snoey E, Jak PMC, Brinkman P, et al. Clinical and inflammatory phenotyping by breathomics in chronic airway diseases irrespective of the diagnostic label. Eur Respir J. 2018;51(1).
2. de Vries R, Muller M, van der Noort V, Theelen W, Schouten RD, Hummelink K, et al. Prediction of response to anti-PD-1 therapy in patients with non-small-cell lung cancer by electronic nose analysis of exhaled breath. Ann Oncol. 2019;30(10):1660-6.
3. Kampf G, Todt D, Pfaender D and Steinmannet D. Persistence of coronaviruses on inanimate surfaces and their inactivation with biocidal agents. Journal of Hospital Infection. 2020. DOI: https://doi.org/10.1016/j.jhin.2020.01.022
4. The central bureau of statistics provides reliable statistical information and data to produce insight into social issues, thus supporting the public debate, policy development and decision-making while contributing to prosperity, well-being and democracy (https://www.cbs.nl/en-gb/about-us/organisation)
5. Fisher RA. The use of multiple measurements in taxonomic problems. Annals of Eugenics, 1936.7(2):p.179-188.
